# Multiple drivers of the COVID-19 spread: role of climate, international mobility, and region-specific conditions

**DOI:** 10.1101/2020.04.20.20072157

**Authors:** Yasuhiro Kubota, Takayuki Shiono, Buntarou Kusumoto, Junichi Fujinuma

## Abstract

The novel Coronavirus Disease 2019 (COVID-19) has spread quickly across the globe. Here, we evaluated the role of climate (temperature and precipitation), region-specific susceptibility (BCG vaccination, malaria infection, and elderly population) and international traveller population (human mobility) in shaping the geographical patterns of COVID-19 cases across 1,055 countries/regions, and examined the sequential shift of multiple drivers of the accumulated cases from December, 2019 to April 12, 2020. The accumulated numbers of COVID-19 cases (per 1 million population) were well explained by a simple regression model. The explanatory power (*R*^2^) of the model increased up to > 70% in April 2020 as the COVID-19 spread progressed. Climate, host mobility, and host susceptibility largely explained the variance of the COVID-19 cases (per 1 million population), and their explanatory power improved as the pandemic progressed; the relative importance of host mobility and host susceptibility have been greater than that of climate. The number of days from outbreak onset showed greater explanatory power in the earlier stages of COVID-19 spread but rapidly lost its influence. Our findings demonstrate that the COVID-19 pandemic is deterministically driven by climate suitability, cross-border human mobility, and region-specific susceptibility. The present distribution of COVID-19 cases has not reached an equilibrium and is changing daily, especially in the Southern Hemisphere. Nevertheless, the present results, based on mapping the spread of COVID-19 and identifying multiple drivers of this outbreak trajectory, may contribute to a better understanding of the COVID-19 disease transmission risk and the measures against long-term epidemic.

## Introduction

The spread of infectious diseases through host–pathogen interaction is fundamentally underpinned by macroecological and biogeographical processes (Murray et al. 2015, 2018; Escobar and Craft 2016); key processes include virus’ origination, dispersal, and evolutional diversification through local transmissions in human societies (Peterson 2014). Since December 2019, Coronavirus Disease 2019 (COVID-19) has quickly spread worldwide from Wuhan, China (Li et al. 2020). The disease transmission geography of COVID-19 is highly heterogeneous; some countries (e.g., Japan) had cases from the earliest stage of this epidemic but had a relatively moderate increase in new cases, whereas others (e.g., EU nations and the USA) have been suffering serious outbreaks. This suggests that the COVID -19 pandemic is driven by complicated factors, which may include habitat suitability, region-specific human mobility, and transmission related to susceptibility. Therefore, identifying the driving force of the outbreak pattern is urgently needed for predicting infection risk at the global scale (Ficetola and Rubolini 2020). Additionally, capturing region-specific factors in relation to the outbreak progress is critically important for improving control measures against the long-term epidemic.

Respiratory virus infectious disease is empirically characterized by a seasonal nature (Altizer et al. 2006). Moriyama et al. (2020) illustrated a framework to better understand the mechanisms of virus transmission; air temperature, absolute/relative humidity, and sunlight are jointly associated with virus viability/stability and host defense, and thereby human-to-human transmission is promoted by contact rates along with host susceptibility (or immunity) to the virus. From this viewpoint, several research groups have focused on relevant factors individually and quickly examined the role of climate (Baker et al. 2020; Araújo and Naimi 2020; Sajadi et al. 2020), international mobility linked to human contact (Wells et al. 2020; Coelho et al. 2020), and community-based host susceptibility (Sala and Miyakawa 2020) in the spread of COVID-19. However, these analyses were inconclusive, and the relative importance of these factors in promoting the disease expansion of COVID-19 remains unclear.

Evaluating the drivers of the COVID-19 spread is a challenging task at the present phase of disease expansion. There is still a distributional disequilibrium in the prevalence of infections globally; the number of confirmed COVID-19 cases changes daily and the trajectories largely differs among countries or regions. Moreover, the absence of population-wide testing for COVID-19 makes it difficult to investigate the growth dynamics of COVID-19 infection. The case data includes selection bias due to surveillance focusing mainly on symptomatic persons. In particular, the availability of the test using reverse transcription polymerase chain reaction (PCR) to identify COVID-19 cases, e.g. the number of PCR tests conducted per population, varies greatly among countries with different medical/public-health conditions (https://ourworldindata.org/covid-testing). Therefore, the true number of the COVID-19 patients and the dynamics of the disease spread are obscured behind the prevalence of asymptomatic carriers (Mizumoto et al. 2020; Nishiura et al. 2020).

Despite the fact that the number of COVID-19 cases may not be sufficiently reliable for conducting epidemiological analyses, such as modelling growth dynamics of the disease, the available data of the confirmed cases can be still informative for implementing containment and/or suppression measures, because the number of the confirmed cases are directly linked to the consumption of medical resources for combatting against the COVID-19 pandemic. Therefore, this study focused on time-series data regarding the number of the COVID-19 cases confirmed from December 2019 to April 2020, as well as country/region-specific variables, e.g. socio-economic conditions and screening effort (number of PCR tests conducted), that potentially affect the number of cases, and then explored multiple drivers of disease expansion by controlling covariates. Specifically, we evaluated the relative importance of climate (temperature and precipitation relevant to habitat suitability for the virus), region-specific susceptibility (BCG vaccination, malaria infection and the relative frequency of elderly citizens hypothesized to be linked to host susceptibility) and international travel (human mobility) in shaping the current geographical patterns of the COVID-19 spread across the globe.

## Material and Methods

### Data

We compiled geographic data on the number of reported COVID-19 cases per day from December, 2019 to April 12, 2020. For 1,055 countries/regions, we collected the number of COVID-19 cases from various sources (see List of data sources for the COVID-19 cases in the end of this preprint). We then calculated the time (in days) since the onset of virus spread that was defined by the date of the first confirmed case in each country or region. We also collected the number of PCR tests conducted from World Health Organization (WHO) (https://ourworldindata.org/covid-testing) to examine the influence of sampling effort on the number of confirmed cases of COVID-19.

For each country or region, we compiled several environmental variables. For mapping cases of COVID-19, the longitude and latitude representing the largest city and area for the country or region were extracted from GADM maps and data (https://gadm.org/index.html). Based on geocoordinates of the cities, we collected the climatic data of mean precipitation (mm month^−1^) and temperature (°C) for January, February and March (WorldClim) using WorldClim version 2.1 climate data (https://www.worldclim.org/data/worldclim21.html) at the resolution of 2.5 arc-minutes grid cells that contained a country or region.

Regarding international travel linked to the disease transmission, we compiled the average number of foreign visitors (per year) for individual countries/regions from the World Tourism Organization (https://www.e-unwto.org/toc/unwtotfb/current). Then, we calculated the relative frequency of foreign visitors per population of each country or region for the analysis.

Regarding region-specific host susceptibility, we collected the following three epidemiological properties: the proportion of people aged over 65 years (elderly population), the number of people infected by malaria (per year), and information regarding bacillus Calmette–Guérin (BCG) vaccination from the WHO (https://www.who.int/malaria/data/en/) and (https://apps.who.int/gho/data/view.main.80500?lang=en) and the BCG Atlas Team (http://www.bcgatlas.org/). We included these attributes to the analyses based on the assumptions that BCG vaccination and/or with recurrent anti-malarial medications are associate with some protection against COVID-19 (Sala and Miyakawa 2020; Adams and Alshaban 2020). Regarding the BCG data, we compiled the following five attributes referring WHO (https://apps.who.int/gho/data/view.main.80500?lang=en) and the BCG Atlas Team (http://www.bcgatlas.org/): i) the number of years since BCG vaccination started (BCG_year); ii) the present situation of BCG vaccination practice (BCG_type) split into all vaccinated, partly vaccinated, one time vaccinated in the past, or never vaccinated; iii) the relative frequency of after-1980 (last 40 years) BCG vaccination for people less than 1 year old (BCG_rate); iv) the number of BCG vaccinations (MultipleBCG) describing countries as never vaccinated their citizens with BCG, vaccinated with BCG only once, vaccinated with BCG multiple times in the past, or vaccinated with BCG multiple times in the present; and v) tuberculosis cases per 1 million people (TB). These BCG-related variables have strong intercorrelation. Therefore, we reduced the dimensions of these variables (BCG_year, BCG_type, BCG_rate, MultipleBCG, and TB) by extracting the first axis of the PCA analysis: the score of the PCA 1 axis was negatively correlated with the five variables, and thus PCA 1 score multiplied by –1 was defined as the BCG vaccination effect.

We also compiled socioeconomic data for each country or region. The population size, population density (per km^2^) (Gridded Population of the World GPW, v4.; https://sedac.ciesin.columbia.edu/data/collection/gpw-v4), gross domestic product (GDP in US dollar), and GDP per person were obtained from national census data (World Development Indicators; https://datacatalog.worldbank.org/dataset/world-development-indicators).

### Statistical analyses

We investigated the relationship between the environmental variables (climatic, host susceptibility, international human mobility, and socioeconomic factors) and the number of COVID-19 cases (per 1 million population) using the two approaches to assure the results robustness: conventional multiple linear regression and random forest, a machine-learning modeling (Breiman 2001), respectively. We separately modeled the accumulated number of COVID-19 cases (per 1 million population) at successive periods from December, 2019 to April 12, 2020.

In the multiple regression analysis, we set the log-scaled cumulative number of COVID-19 cases at a period as the response variable and the climatic factors (mean temperature, squared mean temperature, and log-scaled monthly precipitation), socioeconomic conditions (log-scaled population density and GDP per person), international human mobility (the relative frequency of foreign visitors per population) and region-specific susceptibility (the percentage of people aged ≥ 65 years, the log-scaled relative frequency of people infected by malaria, and the BCG vaccination effect) as explanatory variables.

We also included the time of infection (measured in days) since the first case confirmed at each country or region and the number of tests conducted for COVID-19 (as a measure of sampling effort) as covariates to control the country/region-specific observation biases. In addition, we used the trend surface method to take spatial autocorrelation into account as a covariate: we added the first eigenvector of the geo-distance matrix among the countries or regions, which was computed using geocoordinates of the largest city, as a covariate (Diniz-Filho and Bini 2005). The explanatory power of the model was evaluated by the adjusted coefficient of determination (*R*^2^). We also calculated the relative importance of each explanatory variable in a regression model according to its partial coefficient of determination and determined the predominant variables that explained the variance in the response variables. The statistical significance of each variable was determined by conducting F-test. All of the explanatory variables were standardized to have a mean of zero and a variance of one before these analyses.

In the random forest model, we used the same set of response and explanatory variables, as well as the same covariates. In each run of the random forest analysis, we generated 1,000 regression trees. The model performance was evaluated by the proportional variance explained. We evaluated the relative importance of each explanatory variable based on the increase in the mean squared error when the variable was permutated.

Before these analyses, we tested collinearity between the explanatory variables by calculating the variance inflation factor (VIF); the largest VIF value was 5.28 throughout the period and VIF at April 12, 2020 was 3.35, indicating the absence of multicollinearity in the regression.

To confirm the testing effort bias on the number of confirmed cases, we conducted an additional analysis that accounted for the number of conducted tests (i.e., sampling efforts) in individual countries/regions, as a covariate in the model. Note that this analysis was applied to the data from 128–700 countries/regions, because testing data for many countries is currently unavailable (https://ourworldindata.org/covid-testing).

All analyses were performed with the R environment for statistical computing (R Development Core Team 2012); ‘sf’ package was used for graphics artworks (Pebesma 2018) and ‘randomForest’ package was used for the random forest analysis (Liaw and Wiener 2002).

## Results and Discussion

COVID-19 (as measured by the number of cases per 1 million population) has rapidly spread across the globe after it first appeared in Wuhan, China in December, 2019 (Li et al. 2020) (Fig. 1; Supplement Video 1 https://youtu.be/ZIDMtbek-48), although the epidemic seemed to be in particular biome areas (Fig. 2; Supplement Video 2 https://youtu.be/KlnpUY51D3k). The accumulated numbers of the COVID-19 cases (per 1 million population) according to the progress of disease spread were significantly correlated with the variables related to climate, international human mobility, and host susceptibility, at successive periods since December, 2019 (Fig. 3).

**Fig. 1.**
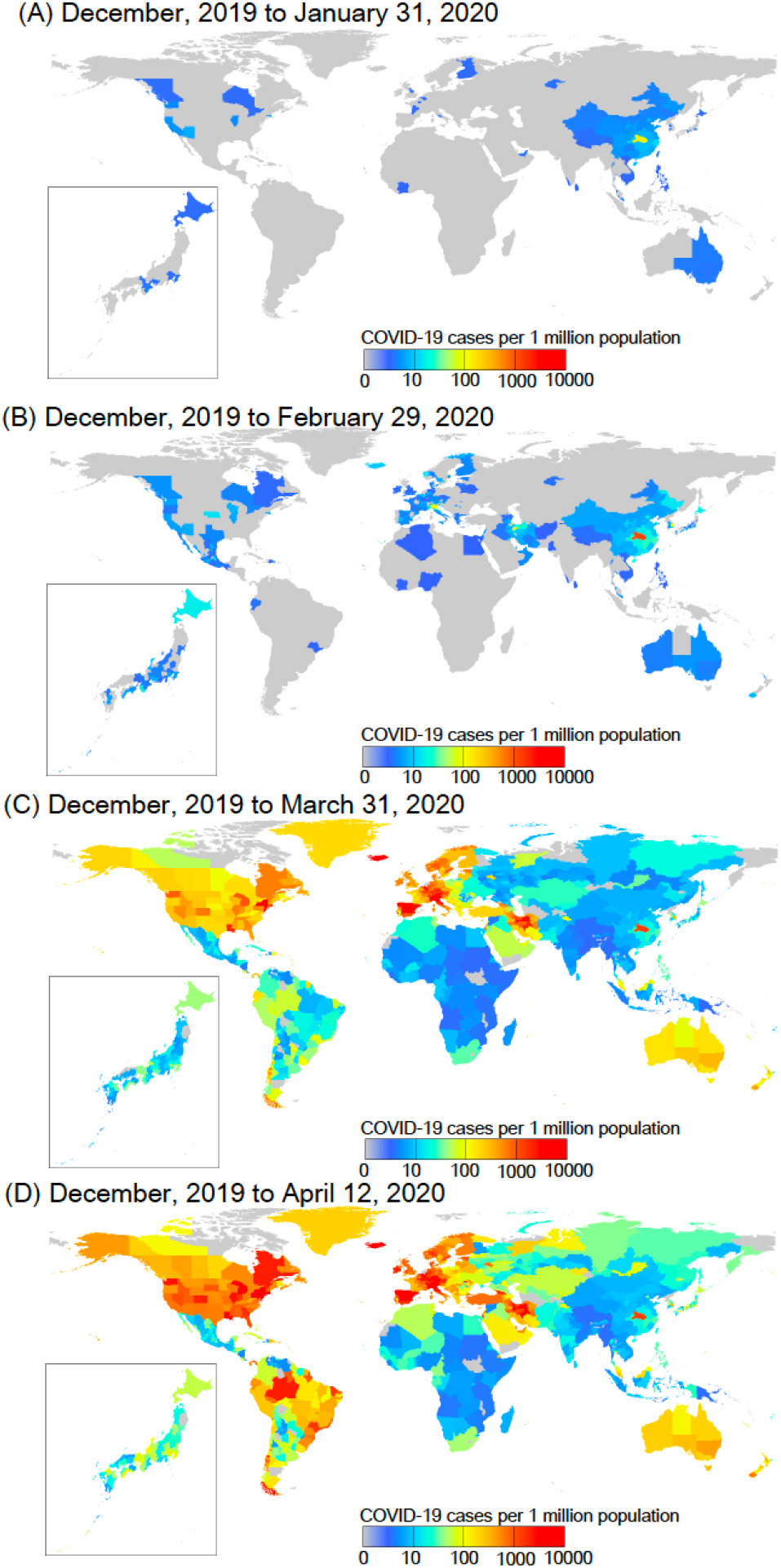
Geographical distribution of COVID-19 cases (per 1 million population) for 1,055 regions worldwide. The monthly patterns of the number of accumulated cases on February 1, 2020 (A), March 1, 2020 (B), April 1, 2020 (C), and April 12, 2020 (D) are based on the accumulated number of day-to-day COVID-19 cases from December, 2019. See Supplement Video 1 https://youtu.be/ZIDMtbek-48.

**Fig. 2.**
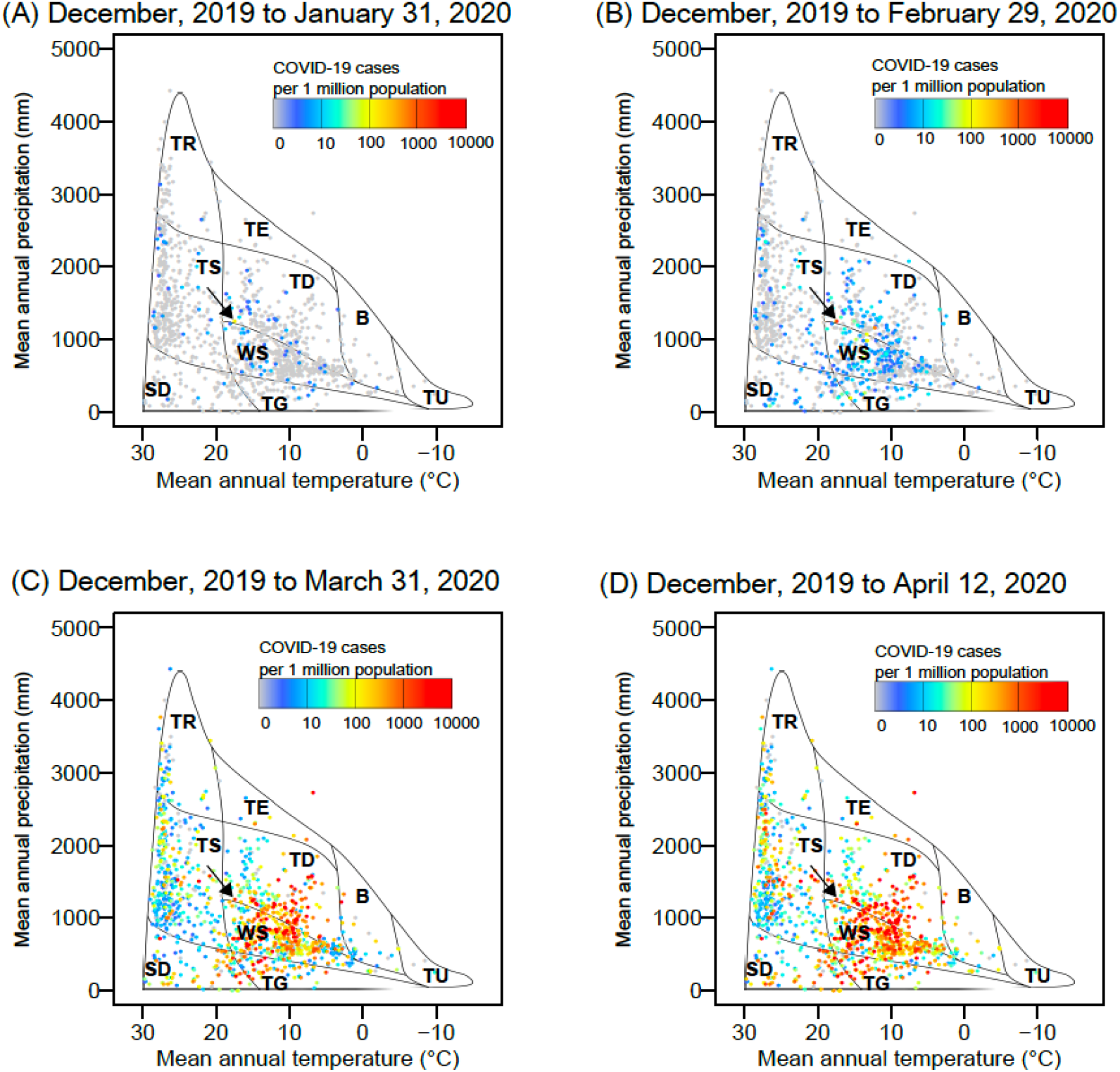
The distribution of the COVID-19 cases across biome types based on the relationship between mean temperature and annual precipitation. Biome classification is based on the scheme by Whittaker (1975). (TR) tropical rain forest; (TS) tropical seasonal forest/savanna; (TE) temperate rain forest; (SD) subtropical desert; (TD) temperate deciduous forest; (WS) woodland/shrubland; (TG) temperate grassland/desert; (TA) taiga; (TU) tundra. Colors indicate the number of COVID-19 cases (per 1 million population). The monthly patterns of the number of accumulated cases on February 1, 2020 (A), March 1, 2020 (B), April 1, 2020 (C), and April 12, 2020 (D) are based on the accumulated number of day-to-day COVID-19 cases from December, 2019. Arrows indicate a location of Wuhan in China. See Supplement Video 2 https://youtu.be/KlnpUY51D3k.

**Fig. 3.**
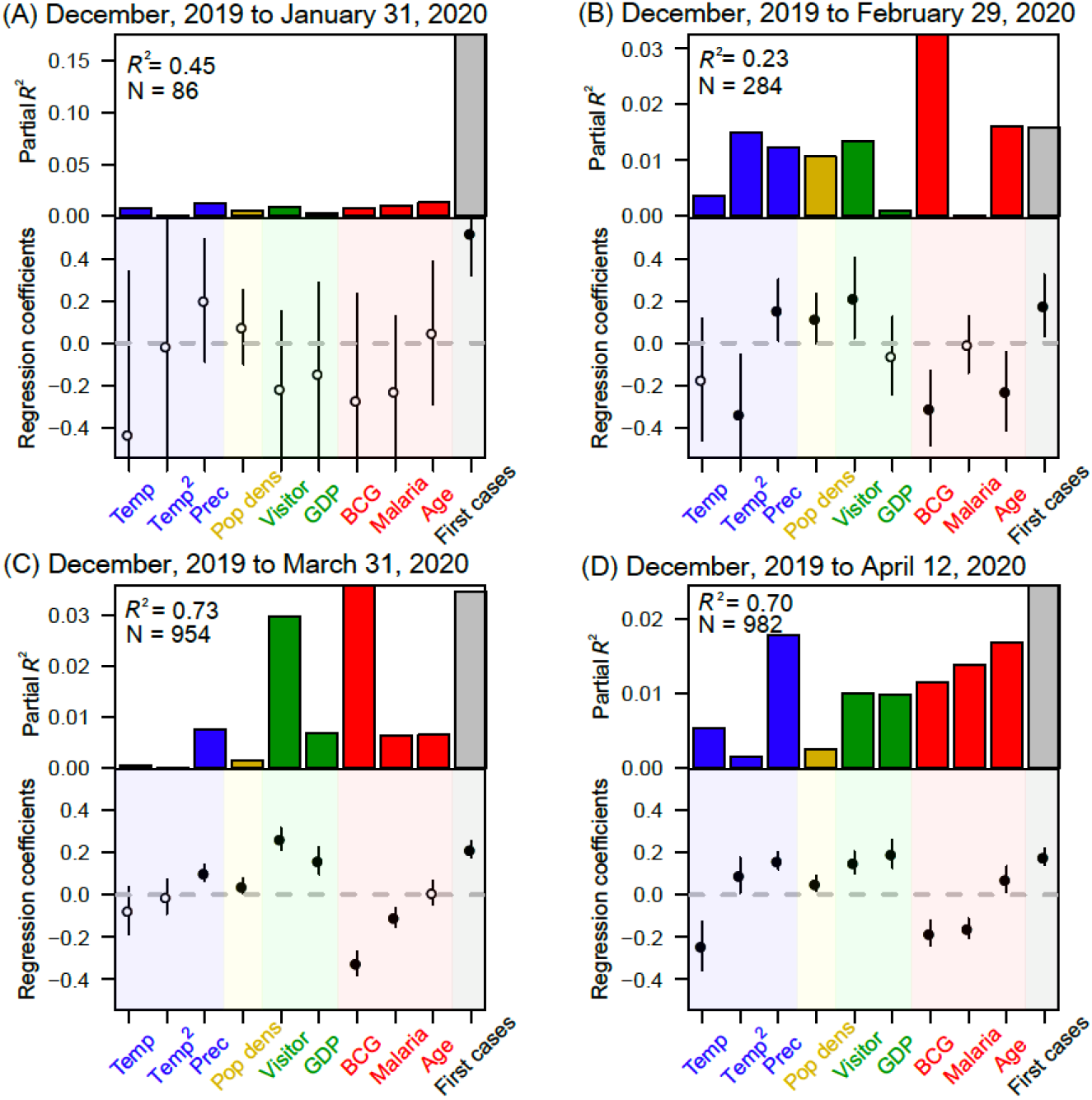
Standardized regression coefficients and coefficient of determination (*R*^2^) of the model explaining the accumulated number of COVID-19 cases (per 1 million population) from December, 2019 to January 31, 2020 (A), December, 2019 to February 29, 2020 (B), December, 2019 to March 31, 2020 (C), and December, 2019 to April 12, 2020 (D). Temp, mean temperature; Temp^2^, squared mean temperature; Prec, mean monthly precipitation; Pop dens, population density; Visitor, relative frequency of foreign visitors per population; GDP, gross domestic product per person; BCG, BCG vaccination effect defined by the first PCA axis summarizing five variables related to BCG vaccination (see the method section); Malaria, relative frequency of people infected by malaria; Age, relative frequency of people ≥ 65 years old; First cases, number of days from case onset. The regressions were conducted using ordinary least squares analyses. Vertical lines represent the 95% confidence intervals of parameters. Closed symbols indicate the significance of explanatory variables (*P* < 0.05). A nonlinear modeling analysis was also conducted by the random forest using the same set of response and explanatory variables and the same covariates; the results of this parallel analysis are shown in Fig. S1.

As time progressed, the standardized regression coefficients of the model greatly changed (from non-significance to significance) through December, 2019 to April 12, 2020 (Fig. 4). After February, 2020, mean temperature was negatively correlated with the accumulated numbers of the COVID-19 cases, whereas mean precipitation was positively correlated (Figs. 4A, B and C). After March, 2020, relative frequency of foreign visitors per population and GDP per person were predominantly positively correlated with the accumulated numbers of the COVID-19 cases (Figs. 4E and F), while BCG vaccination effect and malaria infection were negatively correlated consistently since February or March, 2020 (Figs. 4G and H). Population density was slightly positively correlated (Fig. 4D), and the relative frequency of people ≥ 65 years old was also positively (but temporarily negatively) correlated with the accumulated numbers of the COVID-19 cases (Fig. 4I). In the early stage of COVID-19 spread, the number of days from case onset was strongly positively correlated (Fig. 4J).

**Fig. 4.**
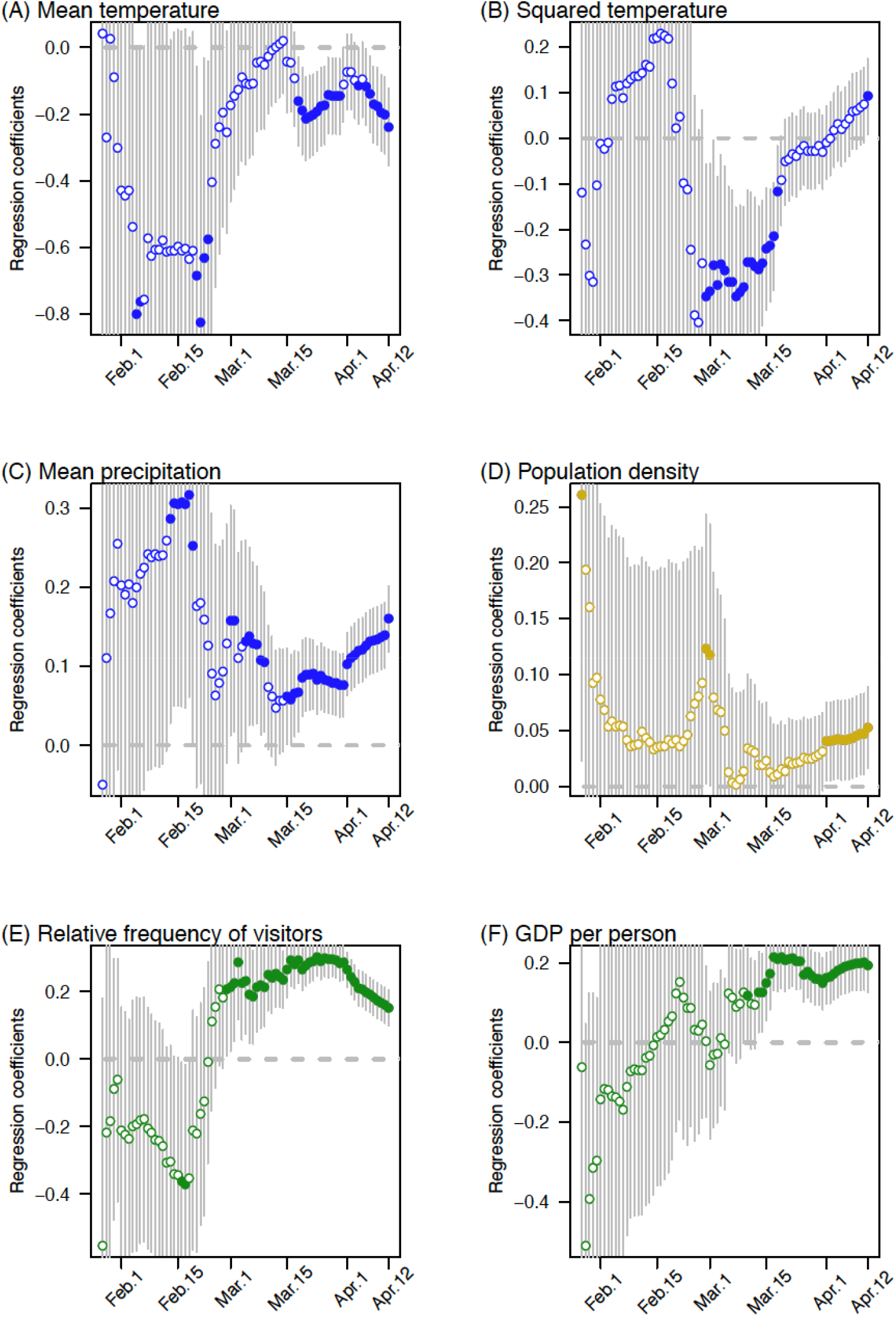
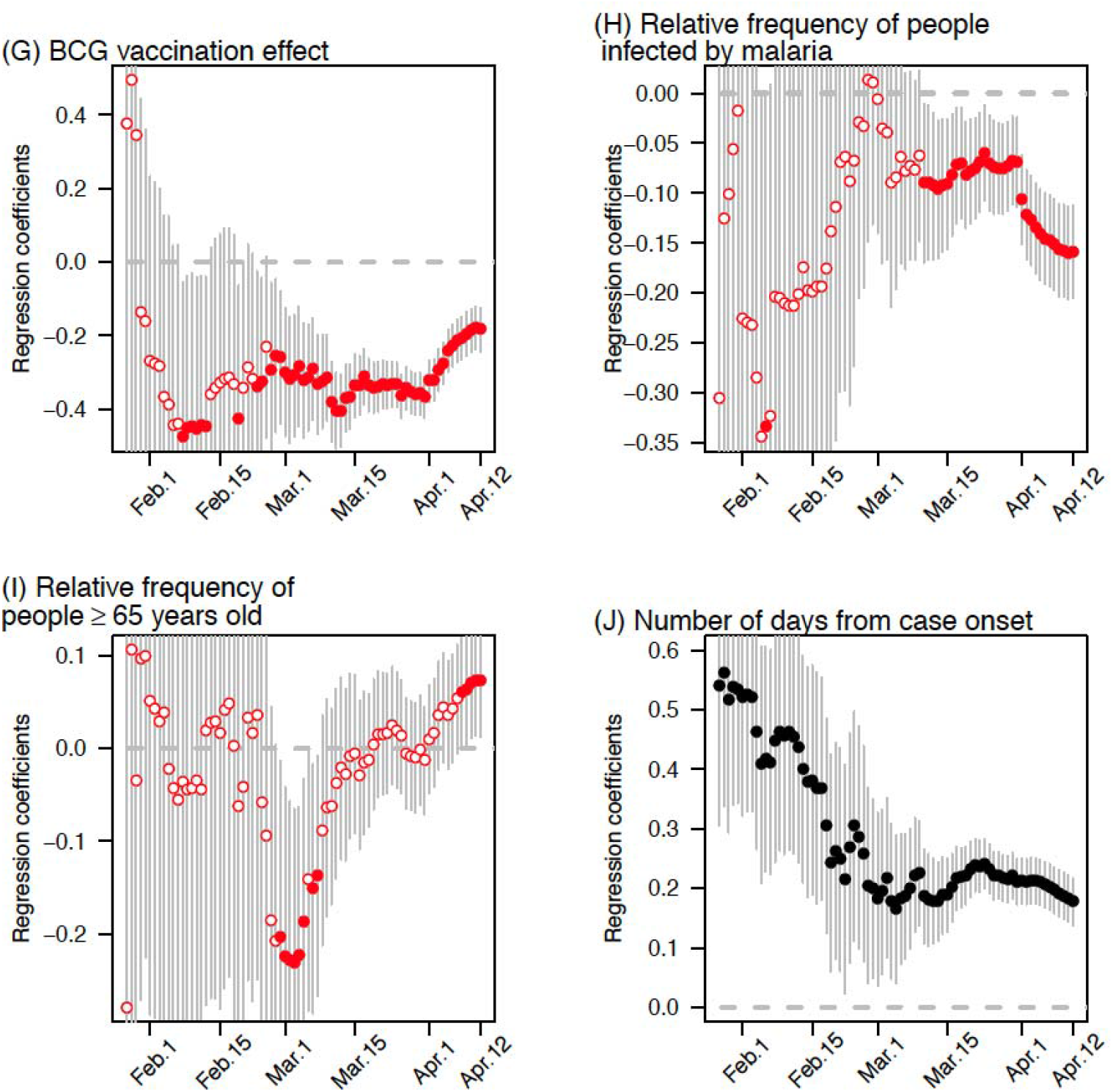
Time-series pattern of standardized regression coefficients (A–J) of the model explaining the accumulated number of the COVID-19 cases (per 1 million population) from December, 2019 to April 12, 2020. The results are shown from January, 2020, because the number of cases in December, 2019 was not sufficient enough for the analysis.

The explanatory power, i.e., coefficient of determination (*R*^2^), of the model increased to up to > 70% in April, 2020 as the COVID-19 epidemic progressed (Fig. 5). The number of days from case onset had greater explanatory power > 20% in January, 2020, but quickly lost its influence as the epidemic progressed, and instead other variables (related to climate, human mobility, and host susceptibility) showed the increasing explanatory powers (Fig. 5).

**Fig. 5.**
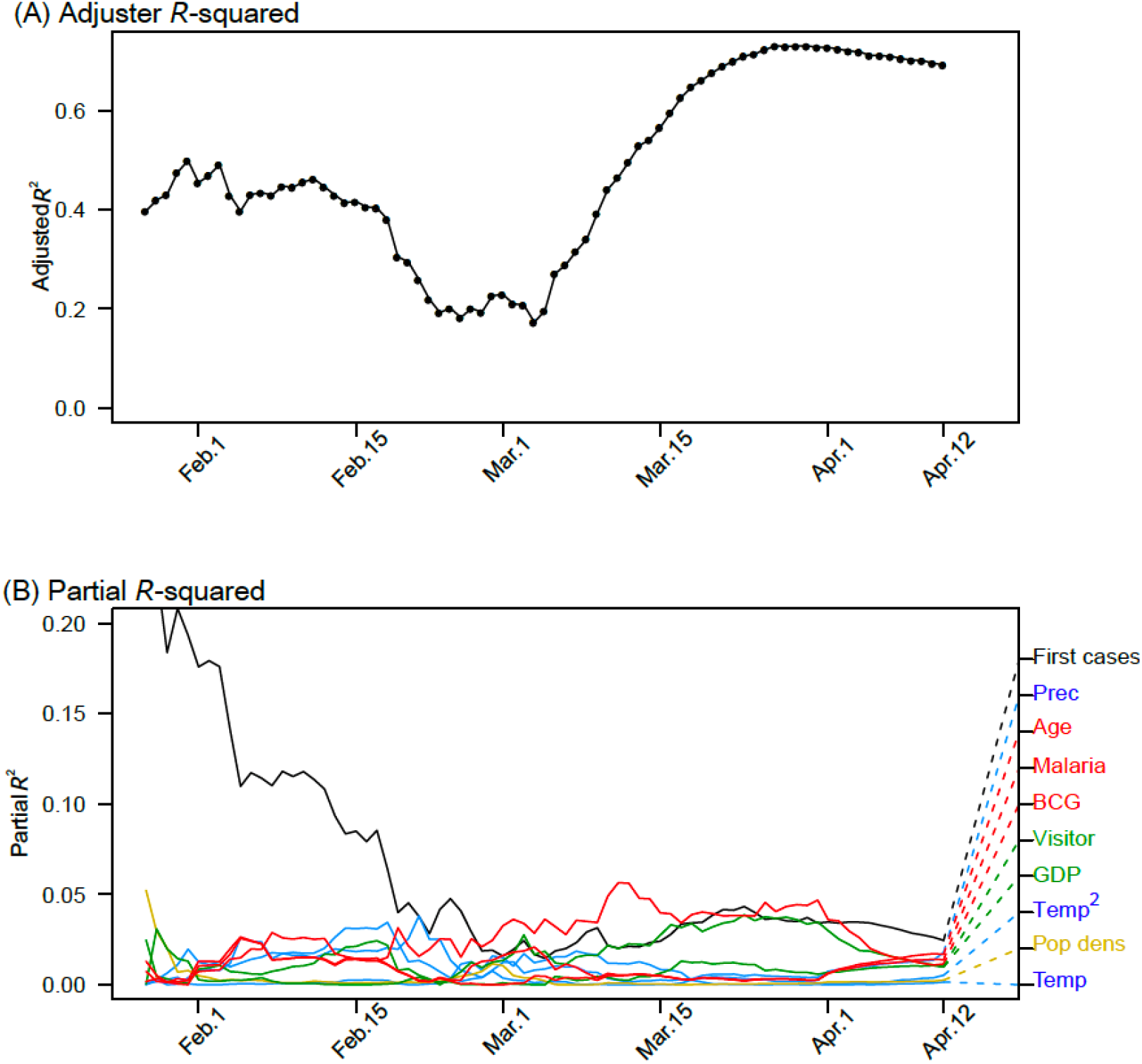
Coefficients of determination (adjusted *R*^2^) of the regression model explaining the accumulated number of the COVID-19 cases (per 1 million population) from December, 2019 to April 12, 2020. (A) Overall coefficient of determination of the regression model; (B) coefficient of partial determination for each explanatory variable in the model. The results are shown from January, 2020, because the number of cases in December, 2019 was not sufficient enough for the analysis.

The results of the random forest model were generally consistent with those of the linear multiple regression model (Fig. S1). The relative importance of the variables related to human mobility and host susceptibility (elderly population, BCG vaccination effect, and malaria infection) became predominant as time progress, whereas that of population density and the number of days from case onset decreased after March, 2020. Moreover, additional analyses, which accounted for the number of tests as a covariate, revealed very similar patterns of regression coefficients of the model and their explanatory power (Fig. S2): the role of climate, international human mobility, and host susceptibility became evident as the epidemic progressed. Therefore, the nonlinearity of epidemic and region-specific testing bias had no serious influence on identifying environmental drivers shaping the present COVID-19 distribution.

This study generally supports the findings of several recent reports, which found that climate (Baker et al. 2020; Araújo and Naimi 2020; Sajadi et al. 2020), international human mobility (Wells et al. 2020; Coelho et al. 2020), and community-based host susceptibility (Sala and Miyakawa 2020) jointly contributed to the spread of COVID-19. Notably, the explanatory power of these drivers substantially increased as the epidemic progressed from January to April, 2020, indicating a deterministic expansion of the disease across the globe.

Cross-border human mobility, which has been facilitated by globalization (Smith et al. 2007), clearly accelerated the COVID-19 pandemic. This finding is in line with a report by Coelho et al. (2020), which emphasized the role of the air transportation network in this epidemic. In addition, region-specific susceptibility, which here was surrogated by BCG vaccination, malaria infection (maybe linked to anti-malarial drugs use), and the elderly population aged over 65 years, explained a substantial part of the variance in COVID-19 cases across the globe: this supports the findings by Sala and Miyakawa (2020) that show a significant correlation between BCG vaccination and COVID-19 prevalence. We note that these correlation patterns may change as this epidemic progresses; a correlation with malaria infection was relatively robust, but the regression coefficients of BCG vaccination effect with the COVID-19 cases (per 1 million population) seems to be less influential after April 2020, which is due to the recent spreads in some particular countries with BCG vaccination program (e.g. Japan, Russia, Turkey and Brazil).

Our analysis using the regression model, which comprehensively accounted for climate, international human mobility, region-specific susceptibility, and socio-economic conditions, showed that climate suitability remains an important driver shaping the current distribution of COVID-19 cases (Ficetola and Rubolini 2020, Araújo and Naimi 2020). Although human mobility and host susceptibility were main drivers in the spread of COVID-19, the distribution of COVID-19 cases across biome types (Fig. 2 and Supplement Video 2) may suggest biogeographical patterns of the epidemic (Murray et al. 2018). Unless the epidemic progresses through the summer months in the Northern Hemisphere and/or in temperate regions of the Southern Hemisphere, it may be too early to make a conclusion on the relationship between abiotic factors and COVID-19 (Moriyama et al. 2020).

Our model prediction does not take into account variables relevant to local-scale factors that are associated with community infection or containment/suppression measures implemented against the epidemic in individual countries/regions. Therefore, the residual of the model (Fig. 6), i.e., deviation of the observed number of COVID-19 cases reflects the influence of local-scale drivers on epidemic spreading; positive deviation may indicate more serious local-scale cluster infection, e.g., in some prefectures in Japan or in parts of South East Asia, Africa, and South America, than we predicted by macro-scale driver-based model, while negative deviation indicates the influence of distributional disequilibrium of COVID-19 cases (because of the areas, e.g., Africa, that the virus reached recently) or suggest the effectiveness of the present control measures.

**Fig. 6.**
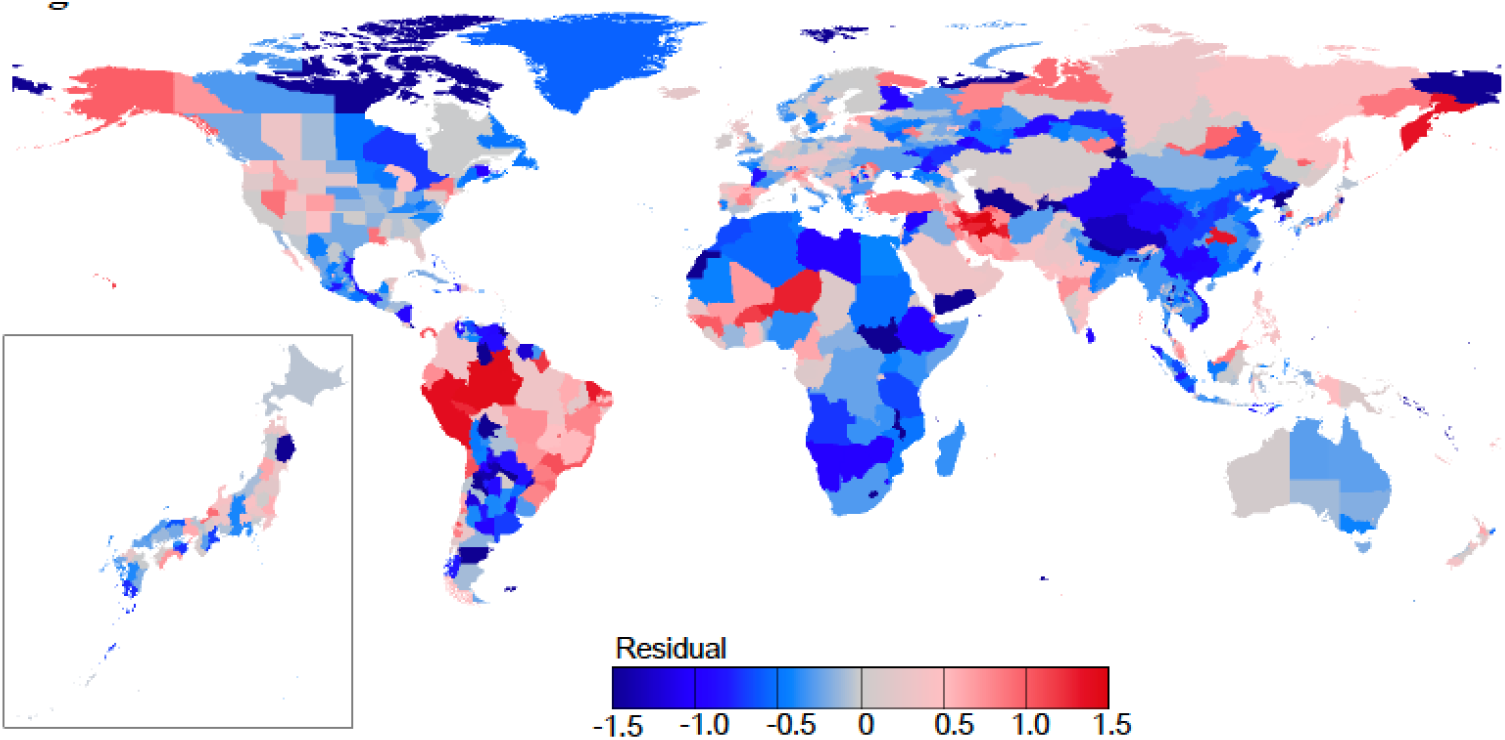
Residual pattern of the regression model predicting the number of COVID-19 cases (per 1 million population) for 1,055 regions across the globe and for 47 prefectures in Japan.

The distribution of COVID-19 cases is not presently at an equilibrium and is changing daily. Nevertheless, our findings demonstrate that the COVID-19 pandemic is deterministically driven by climate suitability, cross-border human mobility, and region-specific susceptibility. The present results, based on mapping the spread of COVID-19 and identifying multiple drivers of this outbreak trajectory, may contribute to a better understanding of the disease transmission risk and the measures against long-term epidemic.

## Data Availability

All data for this study is publicly available. Please see the supplementary material for the source list

## Acknowledgements

We are grateful to the technical staff of Kubota-lab for the data management.

## Author contributions

YK designed the study and wrote the manuscript. TS conducted the data compilation and performed the analysis. KB and JF contributed to the interpretation of study results.

All authors contributed to the final manuscript.

## Notes

### Competing Interest Statement

The authors have declared no competing interest.

### Funding Statement

This study was done voluntarily without public fund.

## References

Adams D. and Alshaban F. (2020) COVID-19 cases are less prevalent in countries where malaria is endemic, suggesting a role for anti-malarial drugs as prophylaxis. Eye Reports 6. doi: http://dx.doi.org/10.16964/er.v6i1.100

Altizer S., Dobson A., Hosseini P., Hudson P., Pascual M., and Rohani P. (2006) Seasonality and the dynamics of infectious diseases. Ecology Letters 9: 467–484.

Araújo M.B. and Naimi B. (2020) Spread of SARS-CoV-2 Coronavirus likely constrained by climate. medRxiv preprint doi: https://doi.org/10.1101/2020.03.12.20034728

Baker R.E., Yang W., Vecchi G.A., Metcalf C.J.E., and Grenfell B.T. (2020) Susceptible supply limits the role of climate in the COVID-19 pandemic. medRxiv preprint doi: https://doi.org/10.1101/2020.04.03.20052787

Breiman, L. (2001). Random forests. Machine learning, 45: 5–32.

Coelho M.T.P., Rodrigues J.F.M., Medina A.M., Scalco P., Terribile L.C., Vilela B., Diniz-Filho J.A.F., and Dobrovolski R. (2020) Exponential phase of covid19 expansion is not driven by climate at global scale. medRxiv preprint. doi: https://doi.org/10.1101/2020.04.02.20050773

Diniz-Filho J.A.F. and Bini L.M. (2005) Modelling geographical patterns in species richness using eigenvector-based spatial filters. Global Ecology and Biogeography 14: 177–185.

Escobar L.E. and Craft M.E. (2016) Advances and limitations of disease biogeography using ecological niche modeling. Frontiers in Microbiology. doi: 10.3389/fmicb.2016.01174

Ficetola G.F. and Rubolini D. (2020) Climate affects global patterns of Covid-19 early outbreak dynamics. medRxiv preprint doi: https://doi.org/10.1101/2020.03.23.20040501

Li Q., Guan X., Wu P., Wang X., Zhou L., Tong Y., Ren R., Leung K.S.M., Lau E.H.Y., Wong J.Y., Xing X., Xiang N. Wu Y., Li C., Chen Q., Li D., Liu T., Zhao J., Liu M., Tu W., Chen C., Jin L., Yang R., Wang Q, Zhou S., Wang R., Liu H., Luo Y., Liu Y., Shao G., Li H., Tao Z., Yang Y., Deng Z., Liu B., Ma Z., Zhang Y., Shi G., Lam T.T.Y., Wu J.T., Gao G.F., Cowling B.J., Yang B., Leung G.M., and Feng Z. (2020) Early transmission dynamics in Wuhan, China, of novel coronavirus–infected pneumonia. N. Engl. J. Med. 382: 1199–1207. doi: 10.1056/NEJMoa2001316

Liaw A. and Wiener, M. (2002) Classification and Regression by randomForest. R News 2: 18–22.

Mizumoto K., Kagaya K., Zarebski A., and Chowell G. (2020) Estimating the asymptomatic proportion of coronavirus disease 2019 (COVID-19) cases on board the Diamond Princess cruise ship, Yokohama, Japan, 2020. Euro Surveill. 12. doi: 10.2807/1560-7917.ES.2020.25.10.2000180

Moriyama M., Hugentobler W.J., and Iwasaki A. (2020) Seasonality of respiratory viral infections. Annu. Rev. Virol. 2020. 7:2.1–2.19. https://doi.org/10.1146/annurev-virology-012420-022445

Murray K.A., Preston N., Allen T., Zambrana-Torrelio C., Hosseini P.R., and Daszak P. (2015) Global biogeography of human infectious diseases. Proceedings of the National Academy of Sciences of the United States of America 112: 12746–12751. https://doi.org/10.1073/pnas.1507442112

Murray, K.A., Olivero J., Roche B., Tiedt S., and Guégan J-F. (2018) Pathogeography: leveraging the biogeography of human infectious diseases for global health management. Ecography 41: 1411–1427.

Nishiura H., Kobayashi T., Yang Y., Hayashi K., Miyama T., Kinoshita R., Linton N.M., JungS-m, Yuan B, Suzuki A., and Akhmetzhanov A.R. (2020) The rate of underascertainment of novel coronavirus (2019-nCoV) infection: estimation using Japanese passengers data on evacuation flights. J. Clin. Med. 9: 419; doi:10.3390/jcm9020419

Pebesma E. (2018) Simple features for R: standardized support for spatial vector data. The R Journal, 10: 439–446.

Peterson A.T. (2014) Mapping disease transmission risk: enriching models using biogeography and ecology. Johns Hopkins University Press, USA.

Sajadi M.M., Habibzadeh P., Vintzileos A., Shokouhi S., Miralles-Wilhelm F., and Amoroso A. (2020) Temperature, humidity, and latitude analysis to predict potential spread and seasonality for COVID-19. Preprint research paper. Electronic copy available at: https://ssrn.com/abstract=3550308

Sala G. and Miyakawa T. (2020) Association of BCG vaccination policy with prevalence and mortality of COVID-19. medRxiv preprint doi: https://doi.org/10.1101/2020.03.30.20048165

Smith K.F., Sax D.F., Gaines S.D., Guernier V., and Guégan J-F. (2007) Globalization of human infectious disease. Ecology 88: 1903–1910. http://dx.doi.org/10.1098/rsif.2014.0950

Wells C.R., Sah P., Moghadas S.M., Pandey A., Shoukat A., Wang Y., Wang Z., Meyers L.A., Singerg B.H., and Galvani A.P. (2020) Impact of international travel and border control measures on the global spread of the novel 2019 coronavirus outbreak. Proceedings of the National Academy of Sciences of the United States of America 117 (13) 7504–7509. doi: https://doi.org/10.1073/pnas.2002616117

Whittaker R.H. (1975) Communities and ecosystems. MacMillan, New York.

